# Climate signal or surveillance artefact? Disentangling environmental and observation processes in Lyme borreliosis trends

**DOI:** 10.64898/2026.09.06.26362392

**Authors:** Maciej Grzybek, Anna Bogacka, Lucinda Kirkpatrick

## Abstract

Climate change is increasingly invoked to explain long-term shifts in vector-borne disease, yet observed incidence reflects both ecological processes and the systems used to detect disease. Distinguishing environmental signals from changes in surveillance is therefore essential for attributing disease trends to global change. We used Lyme borreliosis in Poland as a national case study, analysing 259,003 notifications across 16 administrative regions from 2009 to 2024 (1,024 region-quarters). Multivariable negative binomial models estimated associations with lagged climatic conditions while accounting for temporal trend, a post-2016 structural step, COVID-19 healthcare disruption, seasonality and regional heterogeneity. Hospitalisation proportion provided an independent population-level indicator of changing observed case mix. Reported incidence increased 2.93-fold, whereas hospitalisation proportion declined from 26.4% to 3.5%. Temperature at a three-quarter lag (incidence rate ratio [IRR] 1.069 per 1°C, 95% CI 1.045–1.093) and cloud cover at a one-quarter lag (IRR 1.127 per okta, 1.063–1.195) were independently associated with reported incidence. However, changes in the observation process were of greater magnitude: the post-2016 structural step was associated with a 57% increase in notifications (IRR 1.571, 1.465– 1.684), whereas the COVID-19 period was associated with a 48% reduction (IRR 0.524, 0.482– 0.568). The divergence between increasing notifications and declining hospitalisation indicates substantial expansion in ascertainment of milder disease, such that notification trends cannot be interpreted as proportional changes in transmission. Our findings show that climatic signals can coexist with, and be obscured or amplified by, changes in disease observation. Robust attribution of vector-borne disease responses to global environmental change therefore requires explicit modelling of surveillance processes and integration of epidemiological, healthcare and ecological data.

## 1. Introduction

Detecting biological responses to global environmental change depends not only on the strength of the underlying ecological signal, but also on the stability of the systems used to observe it. This problem is particularly important for vector-borne diseases, in which climatic conditions may influence vector distribution, activity and opportunities for pathogen transmission, while observed human incidence is simultaneously shaped by healthcare access, diagnostic practice, clinical awareness and surveillance sensitivity. Long-term disease-notification series therefore reflect both ecological processes and changes in observation. If these components are not distinguished, temporal trends may be incorrectly attributed to environmental change.

Lyme borreliosis provides an informative system in which to examine this problem. It is the most prevalent vector-borne disease in Europe, with around 132,000 cases notified annually and approximately 30% of the European population living in high-incidence areas [1,2]. Reported incidence has increased across Europe [1], but surveillance systems differ substantially between countries in case definitions, reporting criteria and laboratory-confirmation requirements [3,4]. Within-country changes in diagnostic access, clinical awareness or reporting practice may likewise alter the proportion and clinical spectrum of cases entering national surveillance systems [1,3,5]. Notification streams are therefore generated by healthcare and surveillance systems acting on an underlying ecological transmission process, rather than representing transmission directly.

There is substantial biological plausibility for climatic influences on Lyme borreliosis. Changes in the geographical distribution and environmental suitability of *Ixodes ricinus* have been associated with climatic conditions [9,10], and climate-sensitive indicators of tick-related health risk have increasingly been incorporated into European climate-health assessments [11]. Previous analyses have also identified spatial and temporal associations between Lyme borreliosis incidence and environmental variables, including temperature, precipitation and landscape characteristics [12]. Rising European Lyme borreliosis incidence has therefore frequently been discussed in relation to climate change [1,8]. However, an association between climate and reported incidence does not by itself establish that a long-term increase in notifications represents a proportional increase in transmission. When surveillance sensitivity changes over time, an ecological signal may be amplified, attenuated or obscured by changes in the observation process.

Aggregate notification data alone cannot distinguish an increase in transmission from broader ascertainment of less severe disease. One complementary population-level indicator is the proportion of notified cases that are hospitalised. If the clinical spectrum captured by surveillance remains broadly stable, increasing disease burden would be expected to produce broadly corresponding changes in notified and hospitalised disease. Conversely, a sustained decline in hospitalisation proportion accompanying rising notifications would be consistent with a change in the observed case mix, including progressively greater ascertainment of milder disease. Hospitalisation is not an invariant measure of disease severity, but examining notification and healthcare-utilisation trends together provides an independent means of assessing whether the observation process has remained stable.

Poland provides an informative case study. Reported Lyme borreliosis incidence increased from 27.1 per 100,000 population in 2009 to 79.5 per 100,000 in 2024, placing Poland among Europe’s higher-burden settings [6]. The 16-year time series also contain two pronounced temporal disruptions. Notifications increased from 13,625 cases in 2015 to 21,200 in 2016, a 56% single-year increase with no comparable movement elsewhere in the series, while the COVID-19 pandemic substantially disrupted access to healthcare in Poland during 2020–2021 [7]. Together with sub-national surveillance across all 16 voivodeships and long-term meteorological observations, these features provide an opportunity to examine climatic and observation-related signals within the same analytical framework.

We analysed quarterly Lyme borreliosis surveillance data from all 16 Polish voivodeships between 2009 and 2024, linked to meteorological observations of temperature, cloud cover, precipitation and snow. We aimed to (i) characterise temporal, seasonal and spatial patterns of reported Lyme borreliosis; (ii) identify the best-supported temporal lag for each climatic variable; (iii) estimate climatic associations while accounting for a post-2016 structural shift, the COVID-19 disruption, temporal trend, seasonality and regional heterogeneity; and (iv) evaluate changes in observed case mix using hospitalisation proportion as an independent population-level indicator. More broadly, we asked whether long-term notification data can be interpreted as direct indicators of biological responses to environmental change when the process generating those observations is itself changing.

## 2. Materials and Methods

### 2.1 Study design

We established a regional panel-data framework to examine climatic and observation-related contributions to long-term trends in reported Lyme borreliosis incidence. The unit of analysis was the voivodeship-quarter. The study included all 16 Polish voivodeships and covered 16 consecutive years, from January 2009 through December 2024.

The analytical framework was designed to evaluate whether temporal variation in reported Lyme borreliosis could be explained by climatic conditions alone or whether structural changes in disease observation contributed independently. The time series included two pronounced disruptions relevant to this distinction: a structural step beginning in 2016 and the COVID-19-related healthcare disruption during 2020–2021. Hospitalisation proportion was analysed separately as a population-level indicator of changes in the observed clinical case mix.

### 2.2 Epidemiological data

Lyme borreliosis surveillance data were obtained from annual and quarterly epidemiological reports published by the National Institute of Public Health NIH – National Research Institute (NIZP-PZH-PIB) [6]. Data were available for all 16 voivodeships and included quarterly and annual case counts, incidence per 100,000 population, and numbers of hospitalised cases.

Because the publicly available surveillance dataset was aggregated, individual-level clinical, laboratory, demographic and exposure information was not available. Source files, variable definitions and analytical documentation are provided in the publicly archived research dataset [25].

### 2.3 Climate exposures

Climate observations were obtained from two complementary monitoring networks of the Institute of Meteorology and Water Management – National Research Institute (IMGW-PIB) [13]. The synoptic-climatological (*klimat*) network comprised 28 stations across 12 voivodeships and provided monthly observations of mean air temperature, relative humidity, mean wind speed and total cloud cover. The precipitation (*opad*) network comprised 1,135 stations covering all 16 voivodeships and provided monthly total precipitation, number of days with snowfall and maximum daily precipitation.

Monthly observations were aggregated to quarterly values at voivodeship level. Temperature, relative humidity, wind speed and cloud cover were expressed as quarterly means, whereas precipitation was aggregated as quarterly totals. The station-to-voivodeship crosswalk, intermediate station-quarter data, data dictionary and analytical code documenting the aggregation pipeline are archived in Mendeley Data [25].

### 2.4 Hospitalisation proportion as an indicator of changing observed case mix

Long-term notification series may be affected by changes in the spectrum of disease captured by surveillance. To provide information partly independent of notification counts, we calculated the annual hospitalisation proportion as the number of hospitalised Lyme borreliosis cases divided by the total number of notified cases in each voivodeship-year.

If the clinical spectrum of cases captured by surveillance remained broadly stable over time, substantial increases in notified disease would be expected to be accompanied by broadly corresponding changes in hospitalised disease. Conversely, a sustained decline in hospitalisation proportion alongside increasing notifications would be consistent with a shift in the observed case mix towards less severe disease, potentially resulting from broader case ascertainment.

Hospitalisation proportion was therefore used as an indicator of changing observed case mix rather than as a direct measure of transmission or surveillance sensitivity. Because hospital admission practices may themselves change over time, this analysis was interpreted as complementary evidence rather than as an independent estimate of true infection incidence.

### 2.5 Analytical framework

We used a three-stage analytical framework. Stage 1 characterised temporal, seasonal and spatial patterns in reported Lyme borreliosis incidence. Stage 2 evaluated temporal lags between climatic conditions and reported incidence. For each climate variable, separate negative binomial regression models were fitted at lags of 0–3 quarters. Lag models for a given exposure were fitted to the same fixed sample to ensure comparability, and the best-supported lag was identified using Akaike’s information criterion (AIC) [14].

Stage 3 estimated multivariable models combining climatic and structural predictors. The primary model (Model E) included temperature at lag 3 and cloud cover at lag 1, together with calendar year, quarter, voivodeship, a binary indicator representing the structural step beginning in 2016, and a binary indicator for the COVID-19 period (2020–2021). Population size was included as an offset. Four complementary model specifications were used to assess robustness to climate-variable selection, lag specification and meteorological-network coverage. Full model definitions and results are reported in Supporting Information, Table S2.

### 2.6 Statistical analysis

Quarterly Lyme borreliosis case counts were modelled using negative binomial regression with a log link because of substantial overdispersion in the count data [15]. The logarithm of the population denominator was included as an offset. Models included quarter and voivodeship fixed effects to account for seasonal and time-invariant regional heterogeneity, together with calendar year as a continuous temporal trend. Climatic associations and structural effects are reported as incidence rate ratios (IRRs) with 95% confidence intervals.

The hospitalisation analysis was conducted across 256 voivodeship-year observations. Annual hospitalisation proportion was modelled using ordinary least squares regression including calendar year, annual reported incidence, the COVID-19 period, the structural step beginning in 2016, and voivodeship fixed effects. Regression coefficients are reported with 95% confidence intervals.

Two-sided *p* values were calculated using Wald tests. All analyses were performed in Python 3.12 using NumPy and SciPy; the negative binomial likelihood was implemented directly. The harmonised analytical dataset, statistical analysis plan, analytical code and documentation required to reproduce the analyses are archived in Mendeley Data [25].

### 2.7 Use of artificial intelligence tools

During preparation and revision of this manuscript, the authors used Anthropic Claude Opus 4.7 and OpenAI ChatGPT (GPT-5.6 Sol) to assist with language editing and figure preparation. All AI-assisted output was critically reviewed, edited and verified by the authors against the underlying data and cited literature. The authors retained full control over the study design, data extraction, dataset construction, statistical analyses, model specifications and final scientific interpretation, and take full responsibility for the accuracy, integrity and content of the manuscript.

## 3. Results

### 3.1 Study sample and data coverage

The full surveillance panel comprised 1,024 voivodeship-quarter observations (16 voivodeships × 16 years × 4 quarters) spanning January 2009 to December 2024. Over this period, 259,003 Lyme borreliosis cases were notified nationally. Climate-data coverage varied by exposure network. Temperature, relative humidity, wind speed and cloud cover from the IMGW-PIB *klimat* network were available for 12 voivodeships, yielding 704 voivodeship-quarter observations in the primary analytical sample after lag construction. Precipitation and snow data from the IMGW-PIB *opad* network, comprising 1,135 stations, covered all 16 voivodeships and yielded 944 voivodeship-quarter observations. Four voivodeships (Dolnośląskie, Kujawsko-Pomorskie, Opolskie and Pomorskie) lacked *klimat* station coverage and were therefore included only in analyses based on the *opad* network.

### 3.2 Temporal, seasonal and spatial patterns

Reported Lyme borreliosis notifications increased substantially over the study period, while hospitalisation proportion declined (Fig. 1). National annual case counts increased from 10,332 in 2009 to 29,860 in 2024 (2.89-fold), while reported incidence increased from 27.1 to 79.5 per 100,000 population (2.93-fold). In linear trend analyses, case counts increased by 1,004 cases per year (95% CI 508–1,500; *p*=0.0007), and incidence increased by 2.69 per 100,000 per year (95% CI 1.38–4.00; *p*=0.0006). Over the same period, the national hospitalisation proportion declined from 26.4% in 2009 to 3.5% in 2024.

**Fig. 1.**
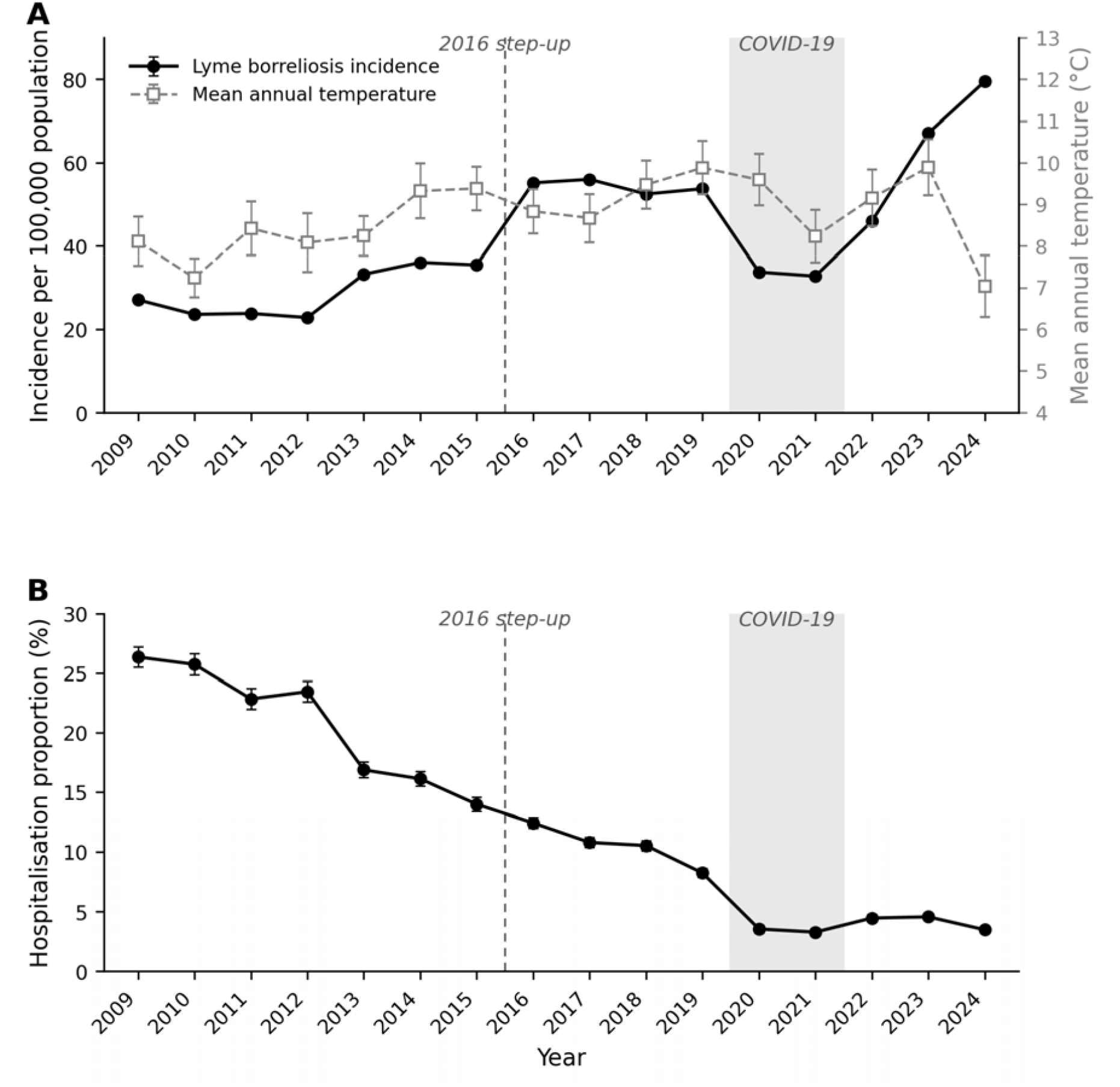
Long-term trends in reported Lyme borreliosis incidence, temperature and hospitalisation proportion in Poland, 2009–2024. (A) Annual national Lyme borreliosis incidence per 100,000 population (black circles, solid line, left axis) and mean annual temperature across climate-covered voivodeships (n=12; grey squares, dashed line, right axis). Error bars for incidence indicate 95% Poisson confidence intervals; error bars for temperature indicate between-voivodeship standard deviation. (B) National hospitalisation proportion (percentage of notified Lyme borreliosis cases hospitalised) by year, with 95% Wilson confidence intervals. The dashed vertical line marks the structural step beginning in 2016; the shaded band marks the COVID-19 period (2020–2021).

A marked discontinuity occurred between 2015 and 2016, when national notifications increased from 13,625 to 21,200 cases (1.56-fold), with no comparable single-year increase elsewhere in the series. Notifications subsequently fell to 12,934 cases in 2020 and 12,500 in 2021 before increasing to 25,285 in 2023 and 29,860 in 2024 (Supporting Information).

A consistent seasonal pattern was observed throughout the study period. Mean quarterly incidence was highest in Q3 (17.75 per 100,000; 95% CI 16.33–19.18), followed by Q4 (13.08; 12.08–14.08), Q2 (8.70; 7.89–9.52) and Q1 (6.03; 5.46–6.60). Spatial heterogeneity was also persistent: mean incidence ranged from 98.9 per 100,000 in Podlaskie and 72.7 in Warmińsko-Mazurskie to 16.9 in Wielkopolskie and 20.8 in Łódzkie, representing an approximately six-fold difference across voivodeships.

### 3.3 Lag structure of climatic associations

Lag-structure analysis identified different best-supported lags across climate variables (Supporting Information, Table S1). Mean air temperature was best supported at a three-quarter lag (IRR 1.064; AIC 7877.2), whereas total cloud cover was best supported at a one-quarter lag (IRR 1.130; AIC 7886.3). Effect estimates for precipitation and snow days were close to unity. Relative humidity and wind speed showed no statistically significant univariate associations. These lag structures informed the primary multivariable specification.

### 3.4 Multivariable associations with reported incidence

In the primary multivariable model (Model E), both climatic and structural variables were independently associated with reported Lyme borreliosis incidence (Table 1).

**Table 1.** Primary multivariable model (Model E) of quarterly reported Lyme borreliosis incidence, Poland, 2009–2024. Negative binomial regression adjusted for quarter, voivodeship, calendar year, COVID-19 period, structural step beginning in 2016, and log population offset (n=704 voivodeship-quarters across 12 voivodeships; AIC=8039.7). Full multivariable comparisons (Models A, B, J and P) are reported in Supporting Information, Table S2.

| Variable | IRR (95% CI) | p value |
| --- | --- | --- |
| Temperature lag 3 (per °C) | 1.069 (1.045–1.093) | <0.0001 |
| Cloud cover lag 1 (per okta) | 1.127 (1.063–1.195) | <0.001 |
| Calendar year (per year) | 1.027 (1.022–1.033) | <0.0001 |
| COVID-19 (2020–2021) | 0.524 (0.482–0.568) | <0.0001 |
| Structural step beginning in 2016 | 1.571 (1.465–1.684) | <0.0001 |

The structural step beginning in 2016 was associated with higher reported incidence (IRR 1.571, 95% CI 1.465–1.684; *p*<0.0001), corresponding to a 57% upward shift after adjustment for climate, seasonality, calendar-year trend and region. The COVID-19 period was associated with lower reported incidence (IRR 0.524, 95% CI 0.482–0.568; *p*<0.0001). A positive residual calendar-year trend remained after inclusion of both structural terms (IRR 1.027 per year, 95% CI 1.022–1.033; *p*<0.0001).

Climatic variables retained independent associations with reported incidence. Each 1°C increase in mean temperature at a three-quarter lag was associated with an IRR of 1.069 (95% CI 1.045–1.093; *p*<0.0001), while each 1-okta increase in total cloud cover at a one-quarter lag was associated with an IRR of 1.127 (95% CI 1.063–1.195; *p*=0.0001). The two climatic predictors were weakly correlated after conditioning on quarter and voivodeship means (*r*=−0.04), with no evidence of problematic multicollinearity.

### 3.5 Sensitivity analyses

Sensitivity analyses showed broadly consistent estimates across alternative model specifications (Fig. 2; Supporting Information, Table S2). In the a priori lag specification (temperature lag 1 and cloud cover lag 1; Model B), cloud cover remained associated with reported incidence (IRR 1.147, 95% CI 1.080–1.219; *p*<0.0001), while the temperature association was weaker (IRR 1.029, 95% CI 1.008–1.050; *p*=0.0075).

**Fig. 2.**
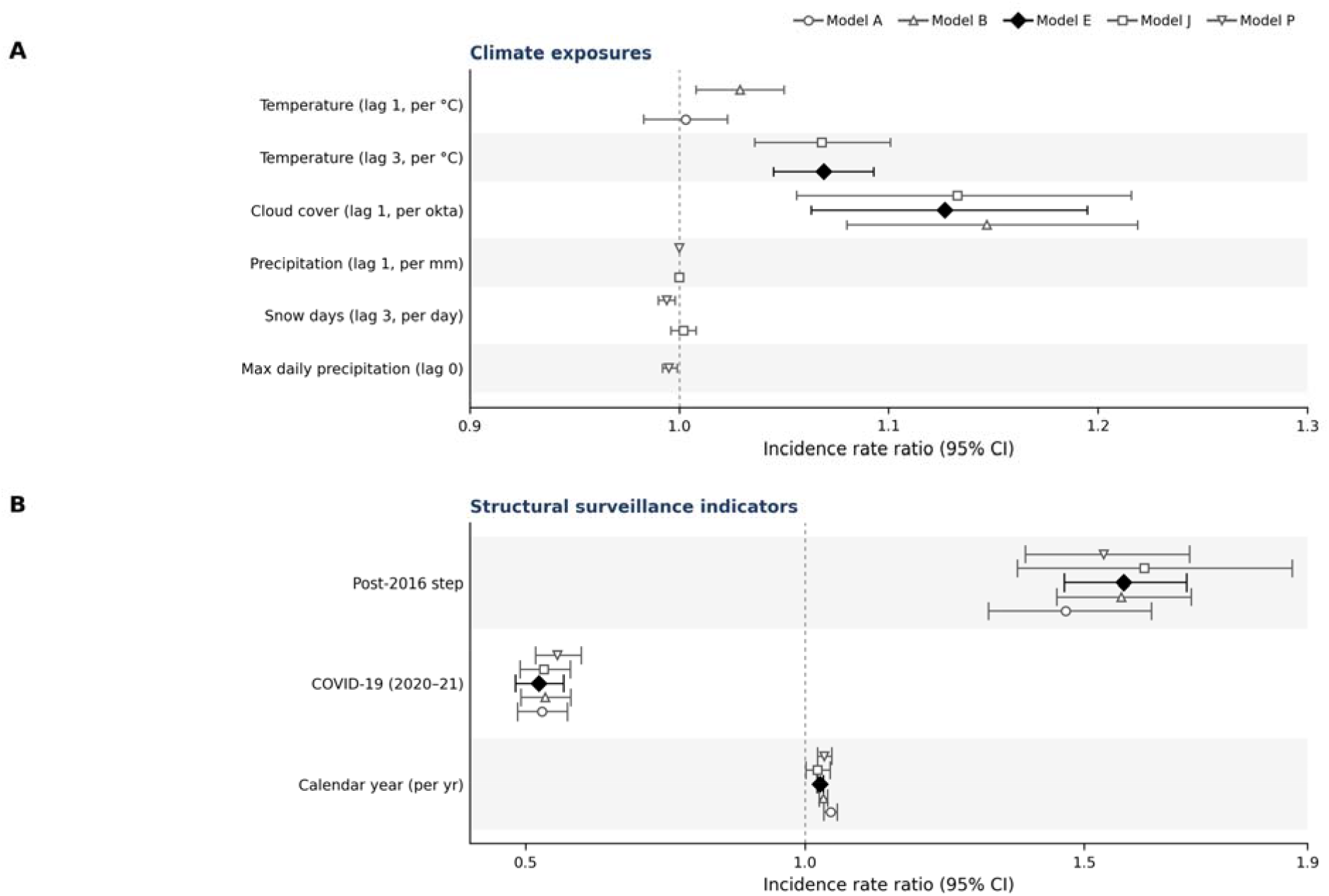
Climatic and structural associations with reported Lyme borreliosis incidence, Poland, 2009–2024. Forest plot showing incidence rate ratios (IRRs) with 95% confidence intervals from the negative binomial regression models. The dotted vertical line indicates IRR=1.00. (A) Climate exposure coefficients; (B) structural-step and COVID-19 coefficients. Marker shapes denote model specifications: circle, Model A; triangle, Model B; diamond, Model E (primary); square, Model J; inverted triangle, Model P.

Adding precipitation and snow days to the primary specification (Model J) did not materially alter the temperature, cloud-cover or structural-step estimates; neither precipitation nor snow days was independently associated with reported incidence in this model. In the full 16-voivodeship precipitation-based model (Model P), estimates for the structural step beginning in 2016 and the COVID-19 period were similar to those obtained in the primary 12-voivodeship model.

### 3.6 Hospitalisation proportion and observed case mix

Across 256 voivodeship-year observations, hospitalisation proportion declined by 1.46 percentage points per year (95% CI −1.75 to −1.18; *p*<0.0001; R^2^=0.77) after adjustment for annual reported incidence, the COVID-19 period, the structural step beginning in 2016, and voivodeship fixed effects.

Annual reported incidence was not independently associated with hospitalisation propor ion (β=−0.006 percentage points per case per 100,000; *p*=0.84). The COVID-19 period was associated with an additional 3.64-percentage-point decline in hospitalisation proportion (95% CI −6.12 to −1.16; *p*=0.004), whereas the structural step beginning in 2016 was not independently associated with hospitalisation proportion (β=−1.80 percentage points; *p*=0.18).

## 4. Discussion

In this nationwide 16-year analysis, reported Lyme borreliosis incidence increased 2.93-fold while the proportion of notified cases requiring hospitalisation declined from 26.4% to 3.5%. At the same time, climatic variables retained independent associations with reported incidence after adjustment for temporal, seasonal, regional and structural factors. Together, these findings indicate that long-term Lyme borreliosis notification trends contain both an environmental signal and substantial variation in the process through which disease is observed. The marked divergence between notifications and hospitalisation proportion suggests that the increase in reported incidence cannot be interpreted as a proportional increase in transmission. More broadly, the findings illustrate a fundamental attribution problem when long-term disease-surveillance records are used as indicators of biological responses to global environmental change.

Rising Lyme borreliosis incidence in Europe has frequently been interpreted in the context of climate change [1,8], supported by evidence of changing climatic suitability for *Ixodes ricinus* activity [11] and by regional associations between reported disease incidence and environmental variables including temperature, precipitation and landscape characteristics [12]. Our results do not contradict an important role for climate. Temperature at a three-quarter lag (IRR 1.069 per 1°C) and cloud cover at a one-quarter lag (IRR 1.127 per okta) remained independently associated with reported incidence after structural and temporal adjustment. Climatic effects are biologically plausible because temperature influences the distribution, phenology and diapause of *Ixodes* ticks [16,17], while warmer conditions have also been associated with increased risk of tick-borne disease [18]. Cloud cover may reflect near-surface moisture, reduced solar radiation or other conditions relevant to tick questing behaviour [17,19]. Precipitation and snow days provided little additional explanatory information after temperature and cloud cover were considered. Thus, climatic variability appears to contribute to the ecological background against which Lyme borreliosis transmission occurs, although the present data do not allow direct estimation of changes in tick abundance, infection prevalence or human exposure.

The central difficulty is that notification incidence represents the end point of a much longer observation pathway. Climatic conditions may affect vector survival, phenology and host–vector encounters, but the subsequent progression from human exposure to recorded disease additionally depends on symptomatic infection, healthcare-seeking behaviour, clinical recognition, diagnostic practice and notification. Consequently, an ecological climate signal may be amplified, attenuated or obscured as it passes through the healthcare and surveillance system. Previous analyses based on notification data necessarily inherit this limitation when the observation process is assumed to remain stable over time. Our findings show why that assumption deserves explicit evaluation: climate associations persisted, but changes associated with the observation system produced shifts in reported incidence of substantial magnitude.

Two pronounced temporal disruptions illustrate this point. The structural step beginning in 2016 was associated with a 57% increase in reported incidence (IRR 1.571) after adjustment for climatic conditions, seasonality, calendar-year trend and regional heterogeneity. The aggregate data cannot identify a single mechanism responsible for this discontinuity, and it should therefore not be attributed to a specific reporting or diagnostic change. Plausible contributing processes include changes in diagnostic access, clinical awareness, healthcare-seeking behaviour, laboratory referral or reporting sensitivity. Heterogeneity between European Lyme borreliosis surveillance systems and discrepancies between surveillance sources have been documented previously [5,20]. The COVID-19 period provided a complementary disruption: reported incidence was almost halved (IRR 0.524), while hospitalisation proportion declined by a further 3.64 percentage points. This pattern is consistent with the documented disruption of healthcare access and Lyme borreliosis reporting during the pandemic [21]. Together, these discontinuities demonstrate that substantial movements in notification incidence can occur independently of gradual environmental change.

The long-term decline in hospitalisation proportion provides an additional perspective on changing disease observation. Across 256 voivodeship-year observations, hospitalisation proportion declined by 1.46 percentage points annually after adjustment for incidence, structural terms and regional effects, while annual incidence itself was not independently associated with hospitalisation proportion. The eight-fold decline in hospitalisation proportion accompanying a nearly three-fold increase in notifications is consistent with a progressive change in the clinical spectrum of disease captured by surveillance, including increased ascertainment of milder cases. However, hospitalisation is not an invariant measure of disease severity. Admission thresholds, outpatient management and clinical practice may also have changed over the 16-year period. Hospitalisation proportion should therefore be interpreted as an independent indicator of changing observed case mix, rather than as a direct measure of surveillance sensitivity or true transmission. Nevertheless, the magnitude and geographical consistency of the divergence provide strong evidence that the observation process did not remain constant through time.

This distinction has implications beyond Lyme borreliosis. Long-term surveillance series are increasingly used to characterise climate-sensitive infectious-disease risk, but notification data are observations generated by coupled ecological and societal systems. If changes in ascertainment are not considered, trends attributed to climate may partly represent changes in the capacity to detect disease. Conversely, the presence of surveillance change does not invalidate genuine climatic associations. The more appropriate interpretation is that both processes can operate simultaneously. For climate-impact assessment, the objective should therefore be not simply to determine whether reported disease correlates with climate, but to establish how much of the observed temporal pattern can reasonably be attributed to biological and environmental processes after accounting for changes in observation.

The study has several strengths. It combines 16 years of quarterly surveillance data from all 16 Polish voivodeships, comprising 1,024 voivodeship-quarter observations and 259,003 notified cases, with observations from two complementary national meteorological networks. The analytical framework explicitly evaluated lagged climatic associations alongside structural discontinuities and included multiple sensitivity specifications. In addition, hospitalisation data provided information on temporal changes in observed case mix that was independent of the climate models. The presence of both the 2016 structural discontinuity and the COVID-19 healthcare disruption within the same time series provided particularly informative contrasts between gradual environmental variability and abrupt changes in disease observation.

Several limitations should also be considered. First, the ecological design precludes individual-level causal inference and does not allow the structural step beginning in 2016 to be partitioned among potential contributing mechanisms. Second, the primary climate-linked analyses were restricted to 12 voivodeships with *klimat* network coverage, although sensitivity analyses using precipitation data from all 16 voivodeships produced similar estimates for the principal structural terms. Third, cloud cover measured in oktas may represent several correlated environmental processes and cannot be interpreted as a direct mechanistic exposure. Fourth, direct ecological measurements, including tick abundance, *Borrelia* prevalence, tick phenology, host-community composition, solar radiation and local microclimate, were unavailable. Age- and sex-specific surveillance data were also unavailable, and residual temporal autocorrelation cannot be excluded. Finally, hospitalisation practice may itself have changed over the study period. These limitations restrict causal interpretation of individual coefficients and prevent direct quantification of true transmission, but they do not alter the observation that reported incidence and hospitalisation proportion followed strongly divergent long-term trajectories.

Future attribution studies would benefit from linking human notification records directly with vector abundance, pathogen prevalence, host ecology and microclimatic observations. Such integration would allow different stages of the climate–vector–host–human observation pathway to be analysed separately rather than inferring ecological change from human notification counts alone. Comparable integration of notification, laboratory and healthcare-utilisation data would also improve interpretation of differences between regions and surveillance systems. One Health surveillance approaches incorporating multiple host groups and pathogen observations offer one route towards such integration [24]. More robust burden estimation is relevant to regional risk assessment [23] and to planning preventive interventions, including candidate Lyme borreliosis vaccination [22].

Long-term notification records remain valuable for detecting changes in vector-borne disease, but they were not designed as direct measures of biological response to climate change. Our analysis demonstrates that significant climatic associations can coexist with large changes in the process through which disease is detected and reported. Treating notification incidence as a direct proxy for transmission therefore risks conflating environmental change with changes in observation. Robust attribution of vector-borne disease responses to global change will require explicit consideration of both processes and, where possible, integration of human surveillance with vector, pathogen, host and environmental data.

## Author Contributions

Maciej Grzybek: Conceptualization; Methodology; Data Curation; Software; Formal Analysis; Visualization; Writing – Original Draft; Writing – Review & Editing. Anna Bogacka: Validation; Writing – Review & Editing. Lucinda Kirkpatrick: Methodology; Writing – Review & Editing. All authors contributed to interpretation of the findings, critically revised the manuscript, approved the final version, and accept responsibility for the work.

## Acknowledgements

We acknowledge the National Institute of Public Health NIH – National Research Institute (NIZP-PZH-PIB) and the Institute of Meteorology and Water Management – National Research Institute (IMGW-PIB) for making their surveillance and meteorological observation data publicly available.

## Ethics Statement

This study used aggregated, routinely collected national surveillance data on reported Lyme borreliosis and publicly available meteorological data. No individual-level, identifiable or sensitive information was accessed, and the study involved no human participants, direct patient contact, interventions, biological sampling or access to medical records. Ethical approval was therefore not required.

Lyme borreliosis surveillance in Poland is conducted within the national legal framework for infectious-disease reporting (Act of 5 December 2008 on the prevention and control of infections and infectious diseases in humans; Journal of Laws 2008 No. 234, item 1570, as amended).

## Data Availability Statement

All data supporting the findings of this study are publicly available. Lyme borreliosis surveillance data are published by the National Institute of Public Health NIH – National Research Institute (NIZP-PZH-PIB) [6], and meteorological observations are available from the Institute of Meteorology and Water Management – National Research Institute (IMGW-PIB) [13]. The harmonised analytical panel, data dictionary, statistical analysis plan, analytical code and documentation required to reproduce the analyses are deposited in Mendeley Data [25] under CC BY 4.0 (data) and MIT (code) licences.

